# Evaluating Cervical Spine Mobility and Fitt’s Law Compliance: The DidRen Laser Test Adapted for Virtual Reality with Age and Sex Effects

**DOI:** 10.1101/2024.06.11.24308762

**Authors:** Frédéric Dierick, Renaud Hage, Wesley Estievenart, Joey Bruno, Olivier Nocent, William Bertucci, Fabien Buisseret

## Abstract

Cervical spine mobility assessment is crucial in rehabilitation to monitor patient progress. This study introduces the DidRen VR test, a virtual reality (VR) adaptation of the conventional DidRen laser test, aimed at evaluating cervical spine mobility.

We conducted a cross-sectional study involving fifty healthy participants that underwent the DidRen VR test. The satisfaction of Fitts’ law within this VR adaptation was examined and we analyzed the effects of age and sex on the sensorimotor performance metrics.

Our findings confirm that Fitts’ law is satisfied, demonstrating a predictable relationship between movement time and the index of difficulty, which suggest that the DidRen VR test can effectively simulate real-world conditions. A clear influence of age and sex on performance was observed, highlighting significant differences in movement efficiency and accuracy across demographics, which may necessitate personalized assessment strategies in clinical rehabilitation practices.

The DidRen VR test presents an effective tool for assessing cervical spine mobility, validated by Fitts’ law. It offers a viable alternative to real-world method, providing precise control over test conditions and enhanced engagement for participants. Since age and sex significantly affect sensorimotor performance, personalized assessments are essential. Further research is recommended to explore the applicability of the DidRen VR test in clinical settings and among patients with neck pain.

## I. INTRODUCTION

Neck pain has a significant prevalence rate, ranging from 22% to 70% within the general population, with approximately 44% of sufferers progressing to chronic symptoms [5]. The implications of neck pain extend beyond discomfort, affecting sensory and emotional experiences and potentially indicating underlying tissue damage in the cervical region. Traditional assessment methods, such as real-world tests have been used to assess the sensorimotor performance of the head-neck complex, providing a better understanding of the pathophysiological mechanisms associated with neck pain, such as the “joint position error” [8] and “The Fly” [28], both of which use a commercially available three-dimensional (3D) electromagnetic motion tracking system. These tests are generally straightforward to implement and affordable to clinicians, both regarding time and budget. The use of sensors, even low-cost ones, to these tests adds complexity but also offers relevant clinical indicators and a means to monitor patient progress and guide rehabilitation efforts [19, 21, 22].

A test of peculiar importance for the present study is the DidRen laser test, with a setup similar to the well-known work of Revel [39, 40], though different in its purpose and complexity. The participants sit on a chair and wears a helmet with a laser pointer. They must perform rapid and precise axial rotations of the head from right to left and back (30*^◦^* from the neutral position, which corresponds to the cervical movements most used during activities of daily living [4]. The laser beam must hit three red visual targets (Figure 1). LEDs directly above the red targets validate each hit in addition to producing a sound [19–23]. The DidRen laser test consists of a simple, standardized, and repetitive sensorimotor task performed in the real world, involving axial head rotations from “target to target” in the same 5-cycles sequence (center–right–center–left–center). In clinical practice, it is particularly useful because it focuses simultaneously on the sensory and motor control systems of the head-neck complex and has many direct neurophysiological connections between the proprioceptive, visual, and vestibular systems [29]. To date, the DidRen laser test has been successfully used to: (1) assess rotatory sensorimotor control of the head at a specific time point [18, 22]; (2) track immediate changes in rotatory sensorimotor control after cervical manual therapy treatment [21]. This test can also be used as a standardized rehabilitation training system.

**FIG. 1:**
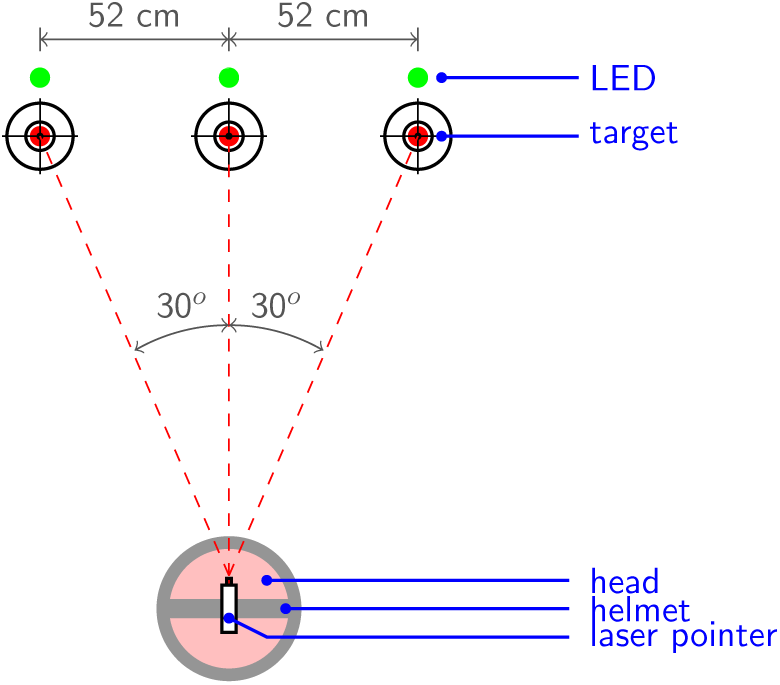
Schematic representation of the real-world DidRen laser test; view from above.

Despite their utility, current assessment methodologies exhibit limitations in adaptability and ecological validity. The fixed and standardized nature of tasks within these tests may not fully capture the dynamic requirements of cervical spine mobility in daily activities. Furthermore, the implementation of sensor-based enhancements, though beneficial, introduces complexities and potential barriers in clinical settings.

The advent of virtual reality (VR) technologies presents an innovative avenue to transcend these limitations. It may provide an interesting framework to implement clinical tests assessing sensorimotor performance of the head-neck complex since: (1) VR headsets are nowadays affordable for all clinicians and contain motion sensors accurately tracking the head motion; (2) VR environments are more modular – more easily customisable – than real-world ones. This may have a positive impact in rehabilitation since the difficulty of the test may be more easily adapted to a given patient and their evolution. An example of such a framework is the “Virtual Reality Test”, developed to assess the functional range of motion of the cervical spine in patients with neck pain [45, 46]. Although this test has a higher sensitivity than the conventional assessment and can also be used to improve the range of motion of the cervical spine [46]. Its major drawbacks relies on the attachment of sensors on the head and the cost for on-site clinical practice.

Similarly, assessing or training a patient with the DidRen laser test in clinical practice is not easy and is very time-consuming, as multiple adjustments must be made to ensure good standardization of the test. Using a VR headset with an environment simulating the DidRen laser test is a promising approach since it allows motion analysis thanks to integrated sensors and the implementation of tests in different spatial directions with vertical (flexion-extension) or even oblique (flexion-extension combined with lateral flexion) movements of the head. Other parameters are in principle easy to modifiy in a VR environment: the distance between the targets, the number of targets, the validation time, the number of cycles, . . . . The VR version of the DidRen laser test, called DidRen VR, has been developed by our team and is presented in Section II.

In view of sensorimotor assessment, the DidRen VR test should not induce different motor strategies than in real-world and therefore ensure that it adheres to established principles of human movement science. Regarding targetting movements, Fitts’ law is a widely accepted model of human locomotor behaviour [13, 14] relying on Shannon’s information theory [47, 48]. In his original investigation, Fitts [13] defined the index of motor performance (*IP*) as

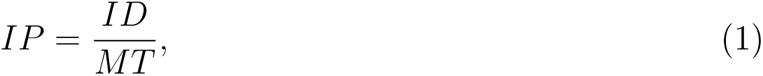

with *ID* the motor index of difficulty and *MT* movement time (*MT*). Note that *IP*, expressed in bits per second, is frequently called throughput in more recent literature. Fitts’ law has been formulated in [14] and states that, in a targetting task, the following link, generalizing (1), holds:

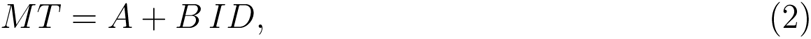

with *A* and *B* empirical positive constants. Moreover, *ID* should depend on the distance between two adjacent target centers (*D*) and on the width (*W*) of the targets through

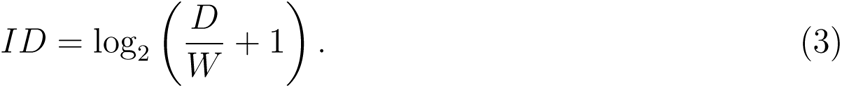

The latter expression for *ID* is not the one originally proposed by Fitts. It is a modified proposal [30, 53] that avoids a negative *ID* for values of *D/W <* 1. Equation (3) directly mimics Shannon’s original theorem #17 [30].

Although Fitts’ law was originally validated for the upper extremities [13, 14], other studies have found that it also describes movements of the lower extremities [11, 54] and head [2, 25]. The mean *IP*, which ranged from 5 [25] to 7 bits*/*s [2], is much lower than values reported for the extremities, which ranged from 10 [13] to 18 [11] or 22 bits*/*s [14]. These studies show that the locomotor system is less efficient in executing pointing movements of the head than that of the extremities for identical values of ID. Therefore, the study of information capacity during the execution of pointing movements of the head, as performed in the DidRen laser test, is of great importance to determine expected values of sensorimotor capacity in healthy subjects or in patients with neck pain.

Turning back to VR applications in motion assessment, clinicians must be able to ensure that there are no conflicts between the real and VR worlds, or that these conflicts are intentional, especially with respect to the position, velocity, and acceleration of different body parts during pointing movements. A simple way to quantify the relevance of a newly developed VR environment and its validity is to calculate Fitts’ law and see if the model is verified. The question we address here is therefore the following: “Does Fitts’ law holds during the DidRen VR test performed in healthy subjects?”. To answer rigorously to this question, the DidRen VR application was designed to allow specific settings for different target widths (challenging the participant’s accuracy) and different inter-target amplitudes (challenging head movement range and velocity), resulting by multiple levels of ID according to Fitts’ law. Our hypothesis is that Fitts’ law will hold during the DidRen VR test. Furthermore, investigating the influence of age and sex on sensorimotor performance within the DidRen VR test is essential. Age-related differences in sensorimotor control and the potential variability in motor performance between sexes could significantly impact test outcomes, paving the way for more personalized and effective rehabilitation strategies. Therefore, the second point of observation was to analyze the effects of age and sex on the VR sensorimotor metrics.

## II. METHODS

### II.1. Participants

The inclusion criteria for the participants were: a Neck Disability Index (NDI) [52] lower than 8% [51, 55], a response to a numeric pain rating scale assessing neck pain equal to 0 – no pain, no neck pain in the last 6 months, no vestibular disorders (vertigo, nausea, motion sickness, etc.), no surgery or trauma related to the cervical spine in the last 6 months, no history of laser eye surgery, no medication that alters perception and no history of epileptic episode(s) or loss of consciousness. Before entering the test phase, each participant completed the French version of the Bournemouth questionnaire [31], which assesses multiple dimensions of neck pain, such as the intensity of pain, associated disability, and its affective and cognitive impacts.

Age and sex of the participants were collected. Participants were divided according to sex into male (M) and female (F) groups.

All the participants were informed about the details of the study through the reading and approval of an informed consent. The study was approved by the ethics Committee of the University of Reims Champagne Ardenne with the approbation number 2022008.

### II.2. DidRen VR test and application

The DidRen VR application, initially developed by one of the co-authors (W.E.) on Samsung Gear VR (SM-R322, Samsung Electronics Co., Seoul, South Korea) and Oculus Quest 1 (Oculus VR Inc., Irvine, CA, USA) in years 2019–2020, was upgraded in 2021 to run on Occulus Quest 2 (Oculus VR Inc., Irvine, CA, USA). This second version of the system is composed of a VR headset and two touch controllers. It is an autonomous LCD display system (dimension [w x h x d]: 91.5×102.0×142.5 mm; mass: 503 g) with price (350– 450 euros), storage (64/256 Gb), resolution per eye (1832×1920), frequency (60/72/90 Hz), field of view (110°), pupillary distance (58/63/68 mm), battery life (2–3 h), and movement tracking possibilities well adapted for clinicians.

The main menu of the application (Figure 2A) allows for managing the application, selecting the desired test, and setting the parameters. A total of 9 tests are available (Figure 3): horizontal (standard spatial setup), vertical, diagonal 1, diagonal 2, triangle up, triangle down, triangle left, triangle right, and random. The accuracy needed to reach a target may be tuned as illustrated in Figure 4. To validate a target, the user has to move the beam by head rotation and stabilize the beam in the validation zone during a particular duration that can be configured. We refer the reader to a video demo, downloadable at https://osf.io/t8zwj/, for a better visualization of the VR environment.

**FIG. 2:**
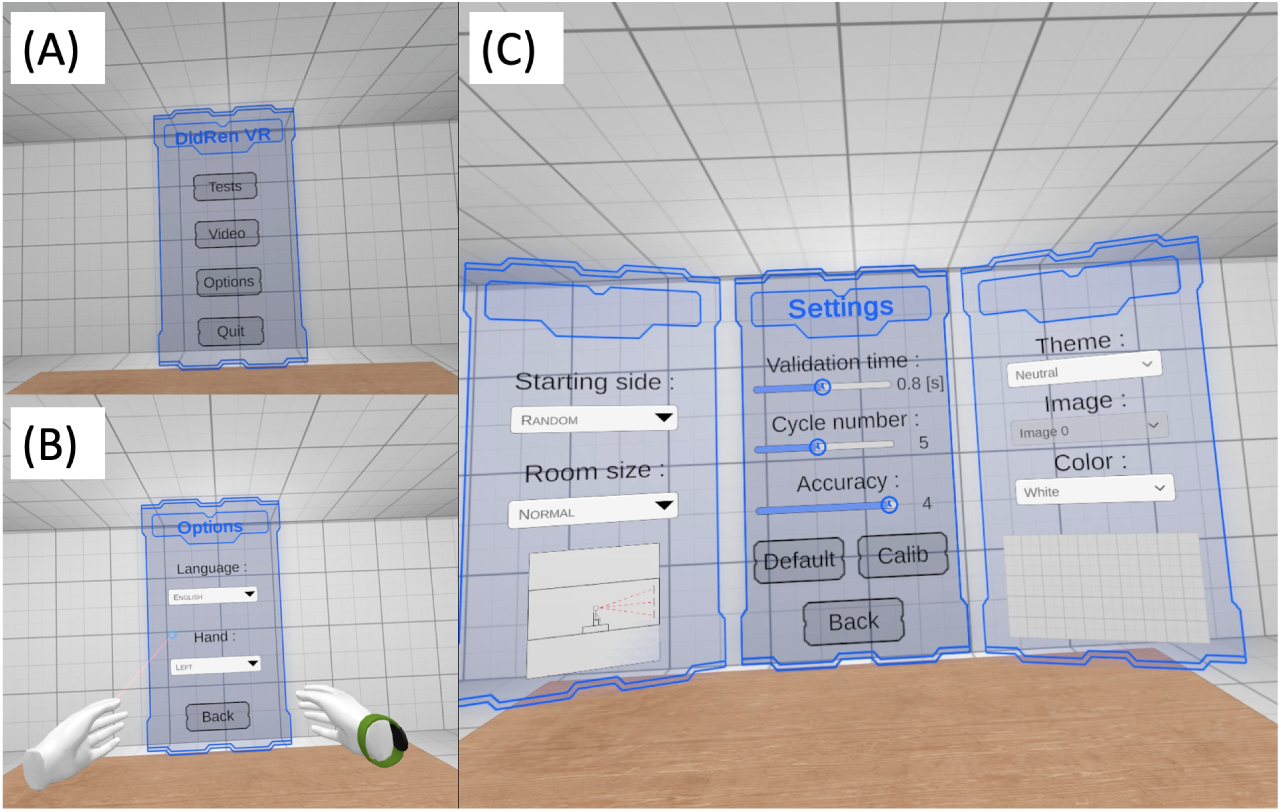
(A) DidRen VR main menu: “Tests” leads to the various environments, “Video” leads to a tutorial, “Options” leads to the choice of language and dominant hand. (B) “Options” menu, allowing to chose language (English, French and Dutch available) and dominant hand. (C) “Tests” main menu allowing to customize the environment. Validation time is the time during which the beam must stay in the validation zone of the target for the target to be considered as reached. Cycle number is the number of cyclic patterns to be performed during the test, e.g. center-right-center-left-center with three horizontal targets. Accuracy is the size of the validation zone of the targets, ranging from 1 (whole target) to 4 (central disk).

**FIG. 3:**
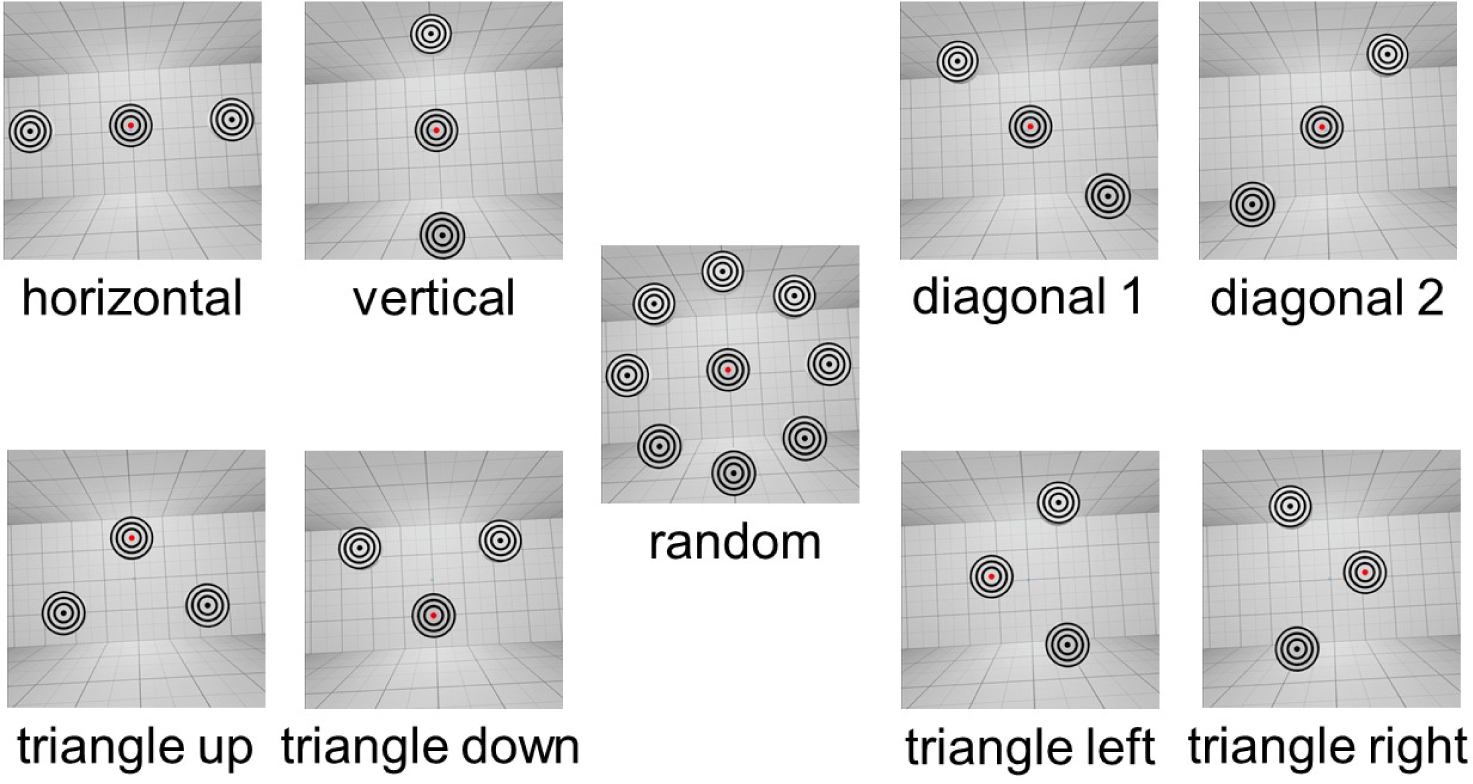
Position of the targets for the 9 developed tests. The horizontal spatial setup corresponds to the standard DidRen laser test. In these illustrations, accuracy is set to 1: The validation zone is the central disk, displayed in red for the target to be reached. Larger accuracy values include the larger concentric circles in the validation zone.

**FIG. 4:**
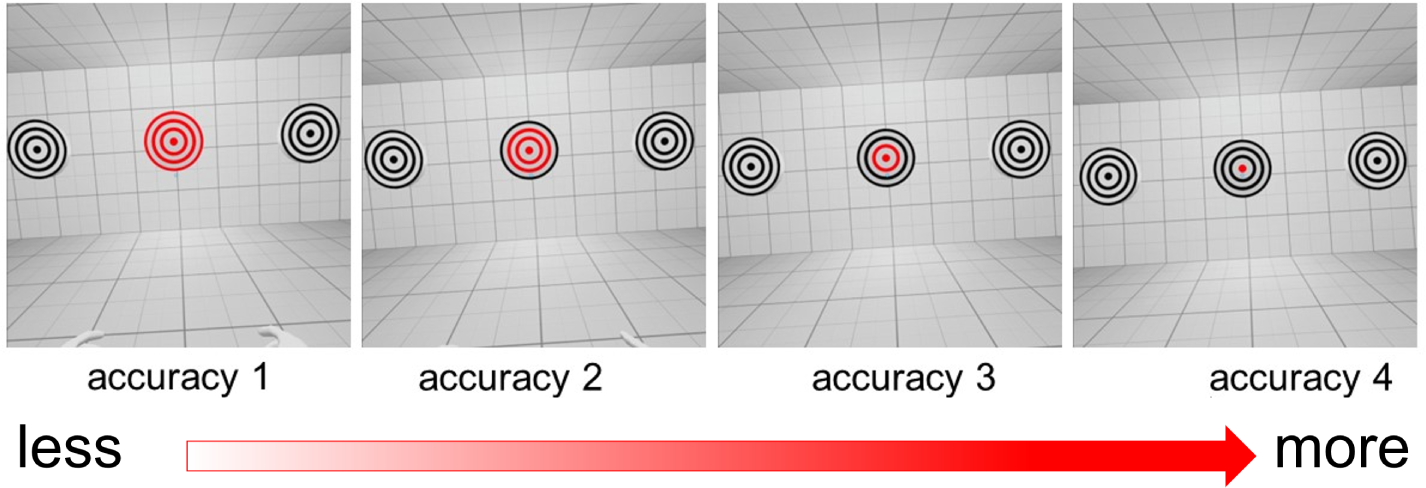
Illustration of the accuracy parameter, ranging from 1 (low accuracy, the whole target is the validation zone) to 4 (high accuracy, the central disk is the validation zone).

In addition to these 9 tests, different parameters can be set directly in the application menu (Figure 2C: the validation time (from 0.1 to 1.5 s by steps of 0.1 s), the number of cycles (from 1 to 10), the size of the validation area or accuracy (from 1 to 4), the starting side (left, right, random), the position of the targets (distance target-floor, distance target-user) and the desired angle (distance between targets). All these parameters are saved and loaded at the next session related to a given participant, so the settings do not have to be changed every time. The positions of the targets can be set in a dedicated menu, and all possible target positions are displayed directly.

During each test, the DidRen VR application records data and creates files that can be used later by the operator. These files contain, on the one hand, the general data, and on the other hand, the 3D kinematic data of the head movements around the X (pitch), Y (yaw), and Z axes (roll) recorded at a sampling frequency of 100 Hz. The general data file includes the identification of the test, the date and the start and end times of the test, the test performed, the room, the total time of the test, the starting rotation side, the target positions, and a link to the corresponding kinematic data file. The latter includes time-series of the following variables: acceleration, velocity, position, angular acceleration, angular velocity (GyrX, GyrY, and GyrZ) and angular position. It is therefore possible to carry out detailed analyses of the movements performed during the test. The application creates the folders, files, and the backup hierarchy itself and checks if the required folders are present before saving/uploading.

The application can be freely downloaded at https://sidequestvr.com/app/9647/didrenvr_demo, where a video presentation can also be found.

### II.3. Experimental procedure

Participants were sat on a chair with a backrest but no armrests and equipped by the experimenter with VR headset (Oculus Quest 2, Oculus VR Inc., Irvine, CA, USA) connected to a computer. Participants were placed in the standard DidRen setup with 3 horizontal targets separated by 30°.

The sequence of head rotations was predefined (centre–right–centre–left–centre, corresponding to one cycle), identical to the classical sequence in real-world DidRen laser test, and similar for all participants that performed five cycles per target distance (angle of rotation). In order to become familiar with the task, a practice test was first carried out before measurements during which instructions were given by the experimenter: *“When a target becomes red you must reach it with the beam as quickly as possible, keep it in the target until it is validated with audible confirmation, then move on to the next one”*.

Participants were then asked to perform the task at 9 different target distances (15° to 55° per step of 5°) in random order. Kinematic data were measured during these 9 tests.

One of the four levels of accuracy was set randomly before the test by the experimenter and the validation time was set equal to 0.8 s instead of the 0.5 s used in the original DidRen test [20]. ID was therefore randomly chosen between a minimal possible value of 0.269 bit for an accuracy level of 1 and an angle of 15°, and a maximum possible value of 3.170 bits for an accuracy level of 4 and an angle of 55°.

At each different angular separation and choice of accuracy, *ID* (Eq. 3) can be computed by equating *D* to the diameter of the validation zone and *L* to the distance between two targets. The movement time, *MT*, is defined by the mean reach time, i.e. the time needed to reach a target that becomes red starting from an other target. This time is computed for each pair of targets and averaged for each pair during the 5 cycles. A linear fit of mean reach time or *MT* versus *ID* is then performed and the *A*- and *B*-values appearing in Eq. (2) are computed, as well as Pearson’s correlation coefficients, *r*. The DidRen VR time (T) and maximal head rotation speed around Y-axis (GyrY_max_) were also collected.

### II.4. Satistical analysis

Data were analyzed using a one-way analysis of covariance (ANCOVA). The dependent variables in this study were *A*, *B*, *r*, T, and GyrY_max_. Age was included as a continuous covariate to account for its potential confounding effects, while sex (M/F) was treated as the independent categorical variable.

Before conducting the ANCOVA, preliminary analyses were performed to ensure the appropriateness of the covariate and the homogeneity of regression slopes assumption. This involved testing the interaction term between age and the sex for each dependent variable. No significant interactions were detected, confirming that the relationship between the covariate and the dependent variables was consistent across levels of the independent variable.

The ANCOVA was conducted separately for each dependent variable. The model for each dependent variable included the main effect of sex, the continuous covariate (age), and the interaction between sex and age, although the latter was subsequently removed from the model if not statistically significant based on a backward elimination approach to maintain model parsimony.

Adjustments for multiple comparisons were made using the Bonferroni correction to control the family-wise error rate. This was particularly important given the multiple dependent variables assessed in our analyses.

Effect sizes were calculated to assess the magnitude of differences observed, which is crucial for interpreting the practical significance of the findings alongside the statistical significance. These were reported as generalized *η* squared (*η*^2^), providing an estimate of how much variance in each dependent variable is accounted for by the independent variable, adjusting for the covariate.

The significance level was set at *α* = 0.05 for all tests. All analyses were performed using R Statistical Software (v4.3.3) [38].

## III. RESULTS

### III.1. Participants

One hundred thirty-seven individuals were recruited and 126 accepted to participate in the study. Seventy-one participants did not meet one or more of the inclusion criteria and five males were excluded for not performing the DidRen VR test correctly (competitive spirit so intense that they did not wait for auditory confirmation of a target before turning their heads to the next one). Finally, 50 participants [28 (56%) males and 22 (44%) females] completed the all protocol and were analyzed (Table I).

**TABLE I:** Description of participants (age, BQ, NDI), results for DidRen VR time at 30° (T) and maximal head rotation speed around Y-axis during DidRen VR test at 30° (GyrY_max_), and results of Fitt’s law parameters (*A*, *B*, and *r*). Data are written under the form median [Q1–Q3] with Q1 and Q3 the first and third quartiles respectively. BQ=neck Bornemouth Questionnaire, NDI=Neck Disability Index.

| Parameter | median [Q1–Q3] |
| --- | --- |
| Age (years) | 39 [23–51] |
| BQ (/70) | 1.5 [0–5] |
| NDI (%) | 0 [0–2] |
| T (s) | 28.9 [27.1–31.1] |
| $\text{GyrY}_{\max}$ ($^\circ s^{-1}$) | 18.0 [11.5–23.7] |
| $A$ (s) | 0.309 [0.214–0.365] |
| $B$ (s bits$^{-1}$) | 0.295 [0.247–0.353] |
| $r$ | 0.938 [0.883–0.968] |

### III.2. Fitts’ law and DidRen VR test

Typical plots of the mean reach time or *MT* versus *ID* data and linear fits between them for three participants are shown in Figure 5. Quality of the fits (or Fitts’ law) could be appraised by the *r* values. The worst (red) and best (blue) linear fits are shown, as well as the median one (green). In all cases it is observed that mean reach time or *MT* versus *ID* follows a clear linear trend. As shown in Table I, the linear fits is clearly followed by the data since the median *r* value was 0.938.

**FIG. 5:**
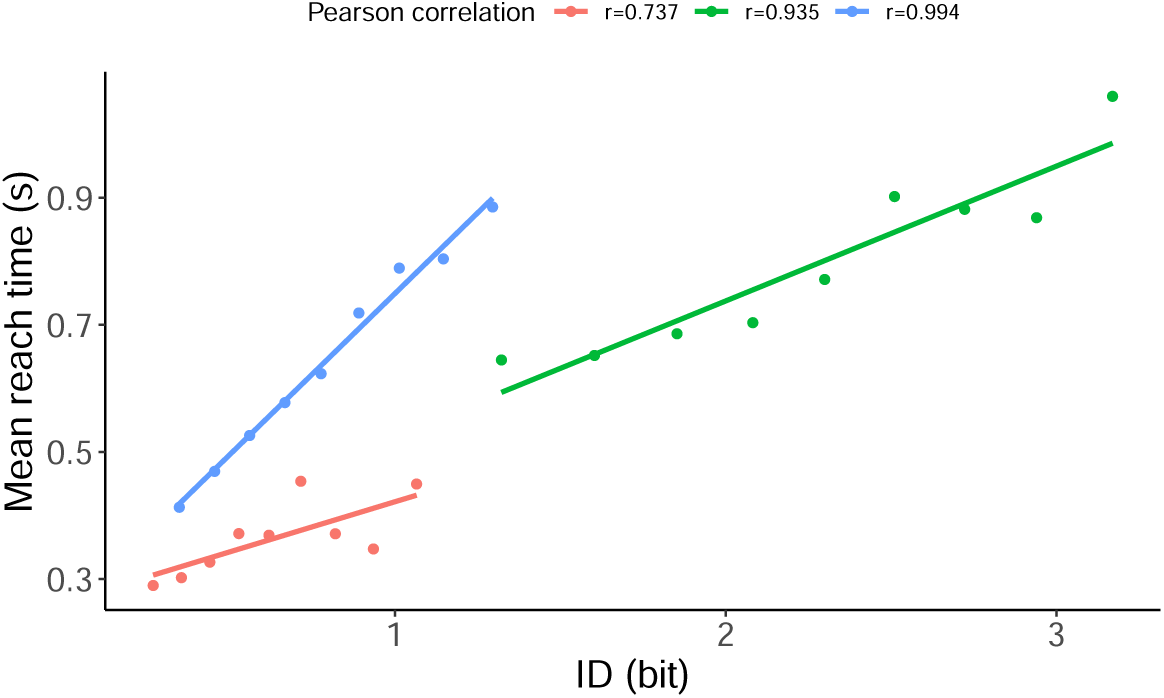
Mean reach time/*MT* (s) versus *ID* (bits) results for three participants. Measured data are plotted (points) and the linear regression corresponding to Fitt’s law has been added (lines). Selected participants are such that their *r* value is minimal (red), maximal (blue), or median (green). Recall that the accuracy level apply during the DidRen VR test was randomly chosen for each participant, leading to different ranges for *ID* values.

### III.3. Age and sex effects

ANCOVA results revealed several significant effects of sex on the dependent variables, after controlling for age. For dependent variable *A*, there was a significant main effect of sex [F(1, 47) = 9.959, *p* = .003, *η*^2^ = .175], indicating that males had lower scores compared to females. Similarly, for variable T, a significant effect was found [F(1, 47) = 6.693, *p* = .013, *η*^2^ = .125], with males showing again lower values than females. *A* and *T* are significantly lower for male than for female participants at any age. ANCOVA results for *A* and *T* are illustrated in Figure 6.

**FIG. 6:**
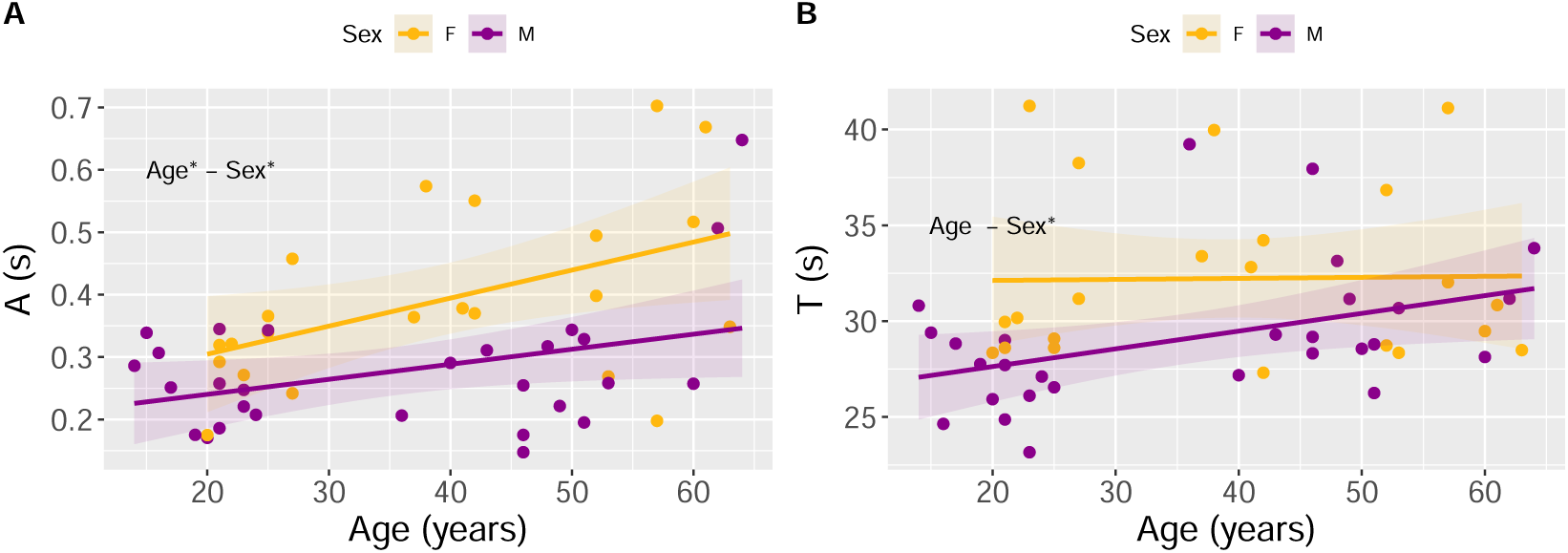
(A) Fitt’s law *A*-value versus participants age. (B) Mean duration of DidRen VR test (T) vs participants age. Males (Females) are displayed in purple (yellow). The linear regressions performed by the ANCOVA are displayed with their 95% confidence interval. * mark significant effect of either age or sex (shifted intercept).

However, no significant effects of sex were observed for variables *B* [F(1, 47) = 0.905, *p* = .346], *r* [F(1, 47) = 0.608, *p* = .439], or GyrYmax [F(1, 47) = 0.853, *p* = .360]. The covariate age, was significantly associated with variable *A* [F(1, 47) = 10.170, *p* = .003, *η*^2^ = .178] but not with other variables, suggesting that age effects were specific to *A*. There were no significant interaction effects between sex and age on any of the dependent variables, confirming that the impact of sex on outcomes was consistent across different ages.

## IV. DISCUSSION

This study introduces the DidRen VR test, an innovative VR adaptation of the traditional DidRen laser test, specifically designed to evaluate cervical spine mobility. The objective of this study was to explore: (1) if the Fitts’ law holds during the DidRen VR test; and (2) the influence of age and sex on various performance metrics.

Our findings confirm that the VR adaptation satisfies Fitt’s law, with correlation values strongly indicating a predictable relationship between movement time and the index of difficulty. This suggests that the DidRen VR test allows for natural cervical movements analogous to real-world conditions and maintains the predictive power of movement time based on target size and distance. Note that, in the real-world DidRen laser test, the index of difficulty is fixed, making impossible a comparison with the present results.

The relationship between movement time and the index of difficulty during the DidRen VR is an other illustration of the linear speed-accuracy trade-off previously described and observed in previous work on the Fitts’ law [10, 13, 36]. Our results were assessed with the time of the DidRen VR test variable which is in line with the previous study of [35], showing similar results.

The results of this study align with the growing body of literature that supports the use of VR in clinical assessments and rehabilitation. Previous research has demonstrated the utility of VR for enhancing engagement and precision in various therapeutic settings [15, 17, 24, 27, 32, 34, 49]. However, our study extends these findings by directly comparing VR adaptations with traditional methods and evaluating their compliance with established motor control theories like Fitt’s law. This compliance is critical, as it underscores the validity of VR platforms in replicating the conditions and challenges of physical tests [12, 43].

Our analysis revealed that age and sex significantly influence performance in the DidRen VR test. It is known from other studies that, although Fitts’ law is satisfied at any age, the latter has an impact on the *A*- and *B*-values appearing in the regression [36, 50]. The fact that Fitts’ law is maintained with age is consistent with age-related deterioration in central processing, planning, or perceptual mechanisms [56]. Using the real laser DidRen test, the elderly have been shown to exhibit altered sensorimotor behaviour, manifested by decreased total time [21]. An obvious outlook of the present study is the assessment of age’s impact on Fitts’ law in the DidRen VR test. According to previous studies [41, 50], the ANCOVA results confirmed the effect of age on *A*-value. *A* reflect the intercept in a plot of movement time against the index of difficulty, suggesting how performance metrics begin or what baseline performance can be expected before difficulty factors are introduced. More, to our knowledge, this is the first time that the effect of sex in relation to head-neck movements has been studied for the Fitts’ law. The ANCOVA results also confirmed the effect of sex, which is very relevant since when studying a population with neck pain, researchers will have to take the slower sensorimotor behavior of women into account.

The implications for rehabilitation are profound, suggesting that VR tests can be tailored to reflect these demographic differences, potentially leading to more customized, effective therapeutic interventions. From an epidemiological point of view, age and female sex are important factors, as the point prevalence of neck pain peaked in middle age, with the highest loads observed in the 45-49 and 50-54 age groups for men and women, respectively [33]. In addition, the point prevalence of neck pain was higher among women in all age groups [26]. And from a clinical point of view, our results are very interesting, since they show that age and sex are already elements that disrupt sensorimotor behavior in pain-free participants. This may mean that the differences that clinicians may note in patients may not be attributable to pain alone, and also that for certain sensorimotor tests no difference may appear between patients and control participants [57].

The median [*Q*1 *− Q*3] time of the Didren VR test at 30° was 28.9 [27.1*−*31.1] s compared to 49.6 [45.6*−*55.6] s in a group of young adults previously studied with the real-world version of the DidRen laser test [20] or to 50.0 [47.1*−*52.8] s for cervicalgic patients aged 46.2*±*16.3 years [21]. The DidRen VR test has therefore a higher index of motor performance than the real-world one: movement time is lower at fixed index of difficulty. It can be hypothesized that a target is more easily validated in the VR environment because there is no loss of accuracy due to laser beam dispersion, dust on the light sensors etc. This may also be the result of an increase in the participants’ engagement due to the VR environment.

Although there is evidence of the efficacy of VR for chronic neck pain [3, 16, 44], the extension of Fitts’s law in VR environments with low-cost technology to assess and train neck mobility is not present in the literature. Our results show for the first time that this law is applicable to the DidRen VR. Therefore, modelling the DidRen in a VR environment does not appear to cause major perceptual conflicts. To avoid these, the targets were placed in a frontal plane (2D) at a perceived distance of 90 cm, as in the real version of the DidRen, although the environment is 3D and the targets can be placed at different distances from the participant. This distance is necessary to control and limit the range of motion of the cervical spine, within a range compatible with use in patients with neck pain. However, using targets at different distances in the DidRen VR would also be a strategy to increase or decrease index of difficulty. In this more complex case, the effect of target depth on cervical mobility performance could be evaluated. A previous study [9] investigated the applicability of Fitts’s law models in 3D environments using the same VR headset as in the present study and showed that target depth affects movement performance. These results suggest that our application should be developed in this direction. The impact of a modification of every parameter has also to be further investigated. For example, the validation time is not adjustable in the real version, so the participant cannot be prompted to reach the next target faster. One strategy that could be used, for example, when assessing patients with neck pain after a training session, is to shorten or lengthen the validation time to avoid training and assessment being similar.

Integrating VR into traditional tests poses several technological challenges, including the accurate tracking of movements, ensuring real-time feedback, and minimizing latency, all of which can affect the test’s efficacy and the user’s experience. Our study addresses these concerns by implementing advanced tracking technologies and optimizing VR software settings, which previous studies have identified as crucial for maintaining test integrity and reliability [1]. It is worth recalling that VR has shown encouraging results in the treatment of neck pain and disability [7, 42]. These studies have shown that patients who used VR improved more than those who did not. However, not all studies show conclusive results [6], so it is important to continue research in this area. Most patients with neck pain do not receive high-intensity, task-oriented training as part of a typical rehabilitation plan. With the DidRen VR, high-intensity repetitive training of neck movements using real and/or misleading ranges of motion, depending on the onset of pain, can now be easily performed in the clinic or at home. In addition, the use of misleading ranges of motion appears to us to be a real option in the treatment of kinesiophobia due to neck pain. This option is currently under development in the DidRen VR application and needs to be tested on neck pain patients in the near future.

While our findings are promising, the study has limitations that need to be addressed. First, the sample size, although adequate for initial analyses, should be expanded to include a more diverse demographic to generalize the findings more broadly. Second, when interpreting the results for intercept A, care must be taken in extrapolating beyond the tested range. Extrapolation involves inherent risks, as it assumes that the established relationships and observed behaviors continue unchanged outside the observed data set. This assumption may not hold if untested variables affect the results, or if the underlying dynamics of the system change beyond the tested thresholds. Thus, while the data within the observed range provide valuable insights, any extension beyond this range should be approached with caution, as it could lead to overgeneralizations that may not accurately reflect real-world scenarios [37]. Third, further studies should explore the long-term effects of using VR for rehabilitation and whether these effects differ from traditional methods in terms of patient outcomes.

## V. CONCLUSIONS

The DidRen VR test presents an effective tool for assessing cervical spine mobility, validated by Fitts’ law, and offers a personalized approach to rehabilitation. The significant effects of age and sex on performance emphasize the need for personalized treatment plans in clinical practices, leveraging VR’s flexibility and engaging nature.

## Data Availability

All data produced are available online at https://osf.io/t8zwj/. Files are described in the README.txt file .

https://osf.io/t8zwj/files/osfstorage/6414602a28e5c50dfb9379db

## VII. ACKNOWLEDGEMENTS

F.D., F.B., R.H. and W.E. acknowledge financial support from the First Haute-Ecole program, project no1610401, DYSKIMOT, in partnership with OMT-Skills (http://omtskills.be/, accessed on 12 March 2022). All authors acknowledge financial support from the European Regional Development Fund: Interreg FWVl 4.7.360 NOMADe (http://nomadeproject.eu, accessed on 01 February 2023).

Data are available here: https://osf.io/t8zwj/. Files are described in the README.txt file . A demo video can be downloaded at https://osf.io/t8zwj/files/osfstorage/6414602a28e5c50dfb9379db.

